# Medical ‘othering’: qualitative evidence synthesis of health services experiences among women affected by female genital mutilation/cutting in Europe

**DOI:** 10.1101/2025.08.13.25333558

**Authors:** Sameera Hassan, Max D López Toledano, Alya Howard, Charnele Nunes, Manar Marzouk, Lam Sze Tung, Oluwafunmilade Adenekan, NUS FGM/C study team, Natasha Howard

**Affiliations:** London School of Hygiene & Tropical Medicine (LSHTM), Faculty of Public Health & Policy, London, United Kingdom; National University of Singapore, Saw Swee Hock School of Public Health, 12 Science Drive 2, Singapore; Faculty of Biology, Medicine and Health, University of Manchester, Oxford Road, Manchester, M13 9PL; University of Nottingham, Division of Psychiatry and Applied Psychology, School of Medicine, United Kingdom

**Keywords:** FGM, FGC, women’s voices, qualitative research, Europe

## Abstract

Migration from countries with high female genital mutilation/cutting (FGM/C) prevalence to Europe has increased attention to health services needs and support for affected women. We explored literature on this foregrounding affected women’s voices, including 37 sources of 11,662 identified. Our findings are synthesised in three themes: medical ‘othering,’ challenged communication, and migration implications. Migrant women affected by FGM/C often struggled navigating health systems that were inflexible to their needs, with health providers often highlighting cultural differences instead of focusing on their health needs in potentially traumatic ways. Women also described feeling torn between conflicting FGM/C cultural norms, their health, and care practices in Europe.

## INTRODUCTION

Public and policy attention to female genital mutilation/cutting (FGM/C) in diaspora communities has increased in Europe, but research remains limited (Baillot et al., 2018). FGM/C is defined by the World Health Organization (WHO) as any “procedure that involves partial or total removal of the external female genitalia, or other injury to the female genital organs for non-medical reasons” (World Health Organization, 2018). It is widely practiced globally, with more than 3 million girls at risk annually, despite many countries - including in Europe - having outlawed it (Mulongo et al., 2014). Increased European immigration from high FGM/C prevalence countries has heightened the importance of understanding the complex effects of FGM/C for affected women and ensuring health services effectively support their needs.

The European Institute for Gender Equality (EIGE) reports that women with FGM/C living in 13 European countries originated from approximately 29 FGM/C practising countries (European Institute for Gender Equality., 2013). Many women living with FGM/C are also immigrants to Europe. Women’s realities as migrants and asylum-seekers include frequent changes of accommodation, general practitioner (GP), and hospitals (Straus et al., 2009). Affected women may feel isolated or unable to discuss FGM/C (Berggren et al., 2006; Vloeberghs et al., 2012). Migration implications alongside potential difficulties navigating new and unfamiliar health systems increase the need for healthcare providers and services that can at least accommodate or ideally holistically support migrant women living with FGM/C in Europe. Poor obstetric outcomes for affected women can relate to living with FGM/C and to migration, thus challenging communication between women and providers, understanding of health information (e.g. due to language barriers), and willingness to accept medical interventions (Vangen et al., 2002). Post-Traumatic Stress Disorder (PTSD), recurrent bad memories, and nightmares can lead affected women to mental healthcare providers (Vloeberghs et al., 2012).

Most literature on health services provision for affected women focuses on healthcare provider perspectives and prevalence estimates of related physical and psychological issues. For example, EIGE estimated the number of girls at risk across 4 European countries as 11-18% from official national datasets (European Institute for Gender Equality., 2013). Studies using qualitative methods emphasise service provider perspectives (e.g. doctors, midwives, allied health professionals) and sexual and reproductive health (SRH). A focus on overall health and support needs of affected women in European settings—from their own perspectives—remains unfulfilled. Hence, this study synthesises the evidence that includes affected women’s descriptions and reflections of their healthcare experiences. As many interventions addressing FGM/C in Europe focus on protecting girls from being cut (Baillot et al., 2018), our approach was to compile narratives of women already affected by FGM/C to better understand how health services could support them. For the purposes of this study, we use the dual term ‘FGM/C’ rather than ‘FGM’ (Female Genital Mutilation) to reduce perpetuation of assumptive ideals regarding the nature and reasoning for this practice rather than to minimise its impact.

This study aimed to explore existing literature that included the voices of women living with FGM/C in Europe on their perceived health needs and experiences, to inform improvements in research and health services provision. Objectives were to: (i) synthesise affected women’s reported needs and experiences engaging with healthcare services in Europe; and (ii) collate potential lessons for health service providers to more holistically support women affected by FGM/C.

## METHODS

### Study design

We collated data from a larger scoping review of FGM/C and female genital cosmetic surgery in Europe and Asia that used Arksey and O’Malley’s six-stage method, Levac *et al*’s 2010 revisions, and Khalil *et al*’s 2016 refinements (Arksey & O’Malley, 2005; Khalil et al., 2016; Levac et al., 2010). Table 1 shows our study definitions.

**Table 1.** Study definitions.

|  |  |
| --- | --- |
| Europe | Greater Europe includes 48 countries (i.e. Albania, Andorra, Armenia, Austria, Azerbaijan, Belarus, Belgium, Bosnia, Bulgaria, Croatia, Cyprus, Czech Republic/Czechia, Denmark, Estonia, Finland, France, Georgia, Germany, Greece, Hungary, Iceland, Ireland/Eire, Italy, Kosovo, Latvia, Liechtenstein, Lithuania, Luxembourg, Macedonia, Malta, Moldova/ Moldavia, Monaco, Montenegro, Netherlands, Norway, Poland, Portugal, Romania, Russia, Serbia, San Marino, Slovakia, Slovenia, Spain, Sweden, Switzerland, United Kingdom) and dependent territories (i.e. Åland Islands, Faroe Islands, Gibraltar, Isle of Man, Svalbard and Jan Mayen) (Kelly et al) . |
| Female genital mutilation/ cutting | Comprises all procedures that involve partial or total removal of the external female genitalia or other injury to the female genital organs for non-medical reasons. The four types are: (i) TYPE 1 or clitoridectomy: partial or total removal of the clitoris; (ii) TYPE 2 or excision: partial or total removal of the clitoris and the labia minora, with or without excision of the labia majora; (iii) TYPE 3 or infibulation: involves narrowing of the vaginal orifice with creation of a covering; (iv) Type 4 includes all other harmful procedures to the female genitalia for non-medical purposes (Baillot et al, 2018). |
| Health system | Consists of all organizations, people and actions whose primary intent is to promote, restore or maintain health, including efforts to influence determinants of health as well as more direct health-improving activities (WHO) (González-Timoneda et al, 2021). |
| Health services | Refers to patient care activities provided through public or private healthcare organizations, by medical professionals and allied healthcare personnel. |
| Voice (as a form of agency) | Voice in this context is 'the capacity to speak and be heard' (World Bank). Agency is the capacity to make decisions about one's own life and act on them to achieve a desired outcome, free of violence, retribution, or fear (Karslen et al, 2020; Mbanya et al, 2020). Voice is not only about giving people the opportunity to communicate ideas and opinions, but should ultimately support their power (agency) to influence change (Karslen et al, 2020). |

### Defining our research question

Our research questions were: (i) ‘What are the scope and main findings of the literature including FGM/C survivors’ voices in Europe?’ and (ii) ‘How do women or girls affected by FGM/C describe their engagement with and experiences of health services and any FGM/C response services?’

### Identifying relevant studies

To increase comprehensiveness, we searched five relevant electronic databases (i.e. CINAHL, EMBASE, Medline[PubMed], ProQuest, PsycINFO) systematically using terms and related terminology for ‘FGM’ and ‘Europe’ adapted to subject headings for each database. We chose EMBASE to access studies focused on obstetrics/gynaecology and healthcare management; MEDLINE and ProQuest for general clinical and public health, also helping compensate EMBASE’s emphasis on English language journals; CINAHL for consumer health and health promotion literature; and PsycINFO for experiences of mental health services. Two university librarians guided database selection and syntax development. Table 2 provides an example for Medline.

**Table 2.** Search syntax and keywords for Medline(PubMed)

| **Controlled vocabulary terms** |  |
| --- | --- |
| Asia | "Asia"[Mesh] |
| Europe | "Europe"[Mesh] |
| FGM/C | "Circumcision, Female"[Mesh] |
| **Keyword statement** |  |
| Asia | "Asia" OR "Afghanistan" OR "Armenia" OR "Azerbaijan" OR "Bahrain" OR "Bangladesh" OR "Bhutan" OR "Brunei" OR "Cambodia" OR "Khmer Republic" OR "Chin*" OR "Cyprus" OR "Georgia" OR "Hong Kong" OR "India*" OR "Indonesia*" OR "Iran" OR "Iraq" OR "Israel" OR "Japan" OR "Jordan" OR "Kazakh*" OR "Kuwait" OR "Kyrgyz*" OR "Kirgizstan" OR "Kirghiz*" OR "Laos" OR "Leban*" OR "Macau" OR "Malaysia" OR "Maldives" OR "Mongolia" OR "Myanma*" OR "Burma" OR "Nepal" OR "Oman" OR "Pakistan" OR "Philippines" OR "Phillipines" OR "Phillippines" OR "Philipines" OR "Qatar" OR "Quatar" OR "Katar" OR "Saudi Arabia" OR "Singapore" OR "Korea" OR "Sri Lanka" OR "Palestin*" OR "West |
|  | Bank" OR "Gaza*" OR "Syria*" OR "Taiwan" OR "Tajikistan" OR "Tadzhik*" OR "Thailand" OR "Siam" OR "Timor-Leste" OR "East Timor" OR "Turkey" OR "Turkmen*" OR "United Arab Emirates" OR "Uzbek*" OR "Vietnam" OR "Viet Nam" OR "Yemen" |
| Europe | "Europe" OR "Åland Islands" OR "Albania" OR "Andorra" OR "Armenia" OR "Austria*" OR "Azerbaijan" OR "Balkan Peninsula" OR "Belarus" OR "Byelorussian*" OR "Belorussian*" OR "Byelarus" OR "Belorussia" OR "Belgium" OR "Bosnia" OR "Bulgaria*" OR "Croatia" OR "Cypr*" OR "Czech*" OR "Czechoslovakia" OR "Denmark" OR "Danish" OR "Estonia*" OR "Faroe Islands" OR "Finland" OR "Finish" OR "France" OR "French" OR "Georgia*" OR "German*" OR "Gibraltar" OR "Greece" OR "Greek" OR "Hungar*" OR "Iceland" OR "Ireland" OR "Irish" OR "Eire" OR "Isle of Man" OR "Ital*" OR "Kosovo" OR "Latvia*" OR "Liechtenstein" OR "Lithuania*" OR "Luxembourg" OR "Luxemborg" OR "Luxemburg" OR "Macedonia" OR "Yugoslav*" OR "Malta" OR "Maltese" OR "Moldavia*" OR "Moldova" OR "Monaco" OR "Montenegro" OR "Netherlands" OR "Dutch" OR "Norway" OR "Poland" OR "Polish" OR "Portug*" OR "Romania*" OR "Rumania" OR "Roumania" OR "Russia*" OR "San Marino" OR "Serbia" OR "Slovak*" OR "Czechoslovakia" OR "Slovenia*" OR "Spain" OR "Spanish" OR "Spanish" OR "Svalbard" OR "Jan Mayen" OR "Swed*" OR "Switzerland" OR "United Kingdom" OR "UK" OR "Engl*" OR "Britain" OR "Scot*" OR "Wales" OR "Welsh" |
| FGM/C | "female genital mutilat*" OR "female circumcis*" OR "clitorectom*" OR "clitoridectom*" OR "infibulat*" OR "genital cut*" OR "ritual female genital surger*" OR "Female W3 genital W3 mutilat*" |
| Female<br>genital<br>cosmetic<br>surgery | ("Labia*" OR "Vagina*" OR "Virgin*" OR "Hymen*" OR "vulva*" OR "genital" OR "hymenoplast*" OR "Labiaplast*" OR "Vaginoplast*" OR "Hymenorrhaphy")<br>AND ("Cosmetic*" OR "Aesthetic*" OR "Restor*" OR "Reduc*" OR "construct" OR "Reconstruct*" OR "Artificial*" OR "rejuvenat" OR "beautif*" OR "tight*" OR "repair")* |

### Selecting studies

Table 3 shows our eligibility criteria, established iteratively based on our research questions. Context was restricted to European countries and topic restricted to FGM/C as defined in Table 1. Outcomes were restricted to affected women’s quotes of their experience. Source types were restricted to academic articles, abstracts, or technical documents. Language was unrestricted for sources with an English abstract. We considered all study designs, interventions, and participants.

**Table 3.** Eligibility criteria.

| Criteria | Included | Excluded |
| --- | --- | --- |
| 1. Context | Greater Europe countries (i.e. Albania, Andorra, Armenia, Austria, Azerbaijan, Balkan Peninsula, Belarus, Belgium, Bosnia, Bulgaria, Croatia, Cyprus, Czechia, Denmark, Estonia, Finland, France, Georgia, Germany, Greece, Hungary, Iceland, Ireland, Italy, Kosovo, Latvia, Liechtenstein, Lithuania, Luxembourg, Macedonia, Malta, Moldova, Monaco, Montenegro, Netherlands, Norway, Poland, Portugal, Romania, Russia, Serbia, San Marino, Slovakia, Slovenia, Spain, Sweden, Switzerland, United Kingdom) or territories (i.e. Åland Islands, Faroe Islands, Gibraltar, Isle of Man, Svalbard, Jan Mayen). | Non-European country or territory. |
| 2. Topic | Any FGM/C. | Studies unrelated to FGM/C. |
| 3. Outcomes | any descriptions about amplifying FGM survivors' voices in relation to health services | Other outcomes. |
| 1. Source type | Primary literature sources (e.g. research-based scholarly journal articles, theses/ dissertations, reports, symposia/ conference abstracts including primary or secondary data).<br>Secondary literature sources (e.g. literature reviews if primary sources are not already included).<br>Commentaries/editorials including primary or secondary data.<br>Reports and book chapters including primary or secondary data. | Tertiary sources with no primary or secondary research data (e.g. encyclopaedias, dictionaries, handbooks, legal/guidance documents).<br>Audio/video reports.<br>Conference abstracts covering the same material as an available publication.<br>Social media, blogs, media articles. |
| 5. Time-period | All | NA |
| 6. Language | All for which an English abstract is accessible. | Sources for which no English abstract is accessible. |
| 7. Study design | Any qualitative | NA |
| 8. Participants | Any | NA |

Figure 1 shows our selection process. First, we identified documents in databases and removed duplicates using the reference manager EndNote. Second, we then screened titles and abstracts against eligibility criteria to remove irrelevant sources using Covidence software. Third, we screened full texts against eligibility criteria to exclude ineligible documents. Finally, we checked for additional documents through backwards-searching reference lists of included studies that we included if eligible.

**Figure 1:**
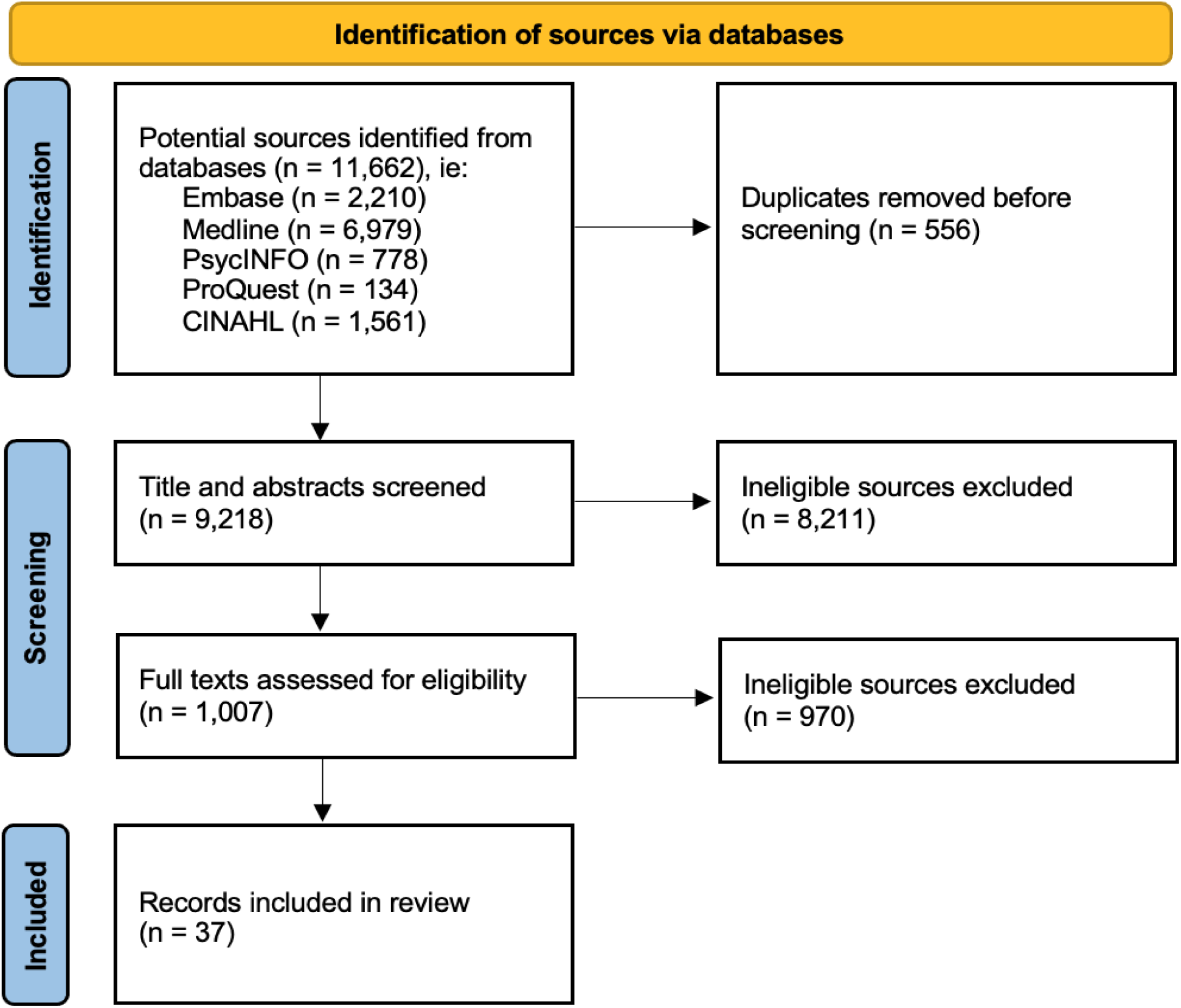
PRISMA Flow Diagram. (Page et al., 2021)

### Extracting data

We extracted data to an Excel sheet using the following headings: (i) source identifiers, i.e. study title, publication year, lead author, source type (i.e. article, conference abstract, report), search type/name (i.e. database [name], reference list); (ii) source characteristics, e.g. country, study design, participant characteristics, time and duration of data collection, study objectives; and (ii) findings, e.g. affected women’s quotes, authors’ related description and analysis.

### Synthesising findings

We summarised source characteristics by extent (i.e. publication year, type), distribution (i.e. publication language, countries included), and nature (i.e. topic/outcomes included, study design, participant characteristics, data collection time-period). We then adopted Noblit and Hare’s seven-step meta-ethnography process for data synthesis (Britten et al., 2002). Similar to thematic synthesis, which seeks to build understanding of various literatures through identifying recurring themes across sources (Kelly et al., n.d.), meta-ethnography goes further by translating concepts and metaphors across studies to generate new conceptualisations or theory, making it more interpretive and, in this case, allowing an ‘ethnographic’ approach to working with women’s perspectives without collecting additional primary data.

## FINDINGS

### Source characteristics

We included 37 qualitative studies of 11,662 identified (Figure 1). Table 4 shows most (12 each) sources described women’s experiences in Norway and UK, followed by 5 in Sweden, 4 in Spain, 2 each in Belgium and the Netherlands, and 1 each for France, Italy, Portugal, and Switzerland. Most (26) included interview or focus group (7) methods, while 4 included participant observation, and 1 each ethnography and arts-based workshop. Sample sizes were generally small, averaging 22 (range 2-70). All studies were conducted by researchers from academic institutions, though some also employed community-led methods. Study participants ranged in age from 13-73 and were either recruited from clinics they attended, or through snowballing within lay settings (e.g. refugee centres, women’s organisations). Health service types included maternal and general or specialist reproductive health services (e.g. re-infibulation).

**Table 4.**
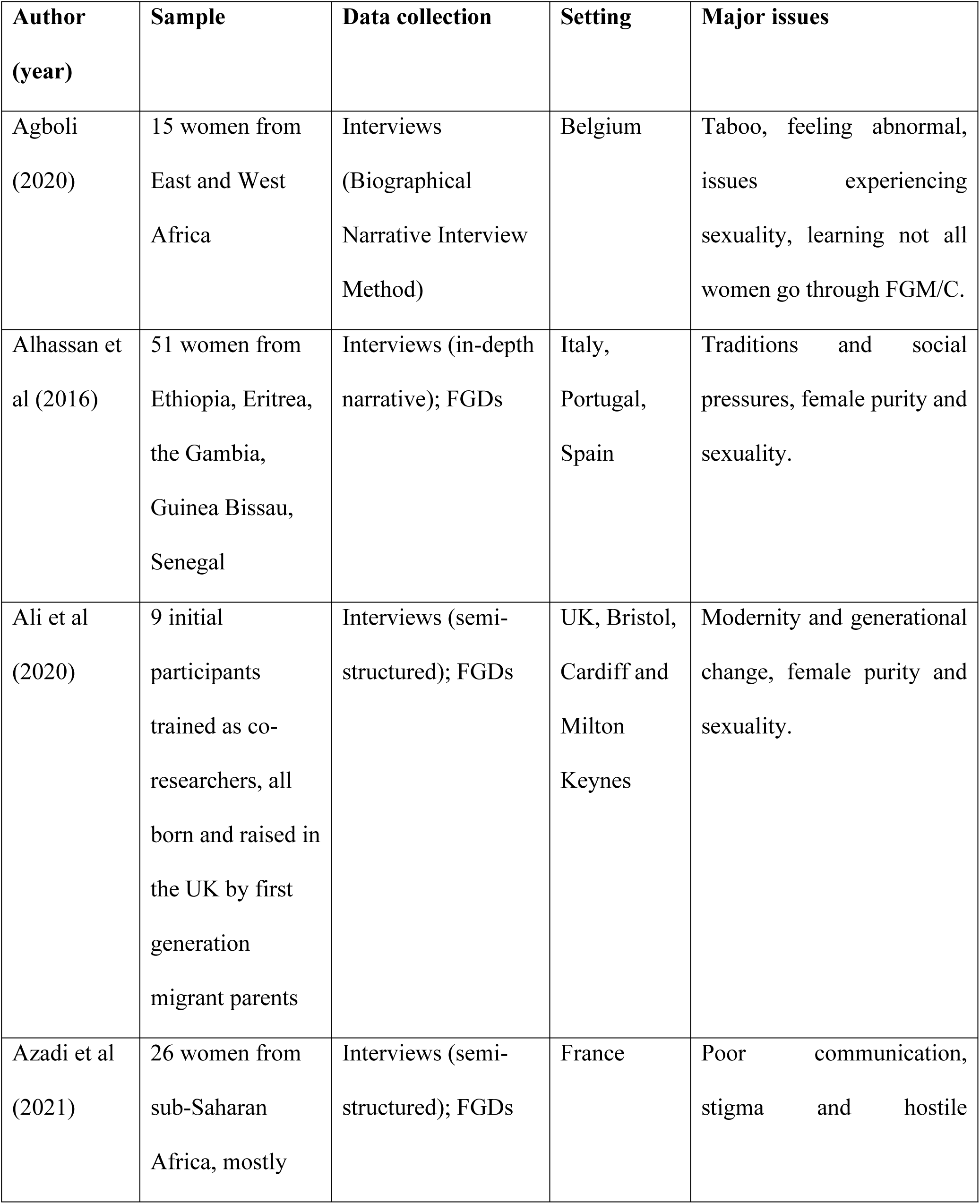
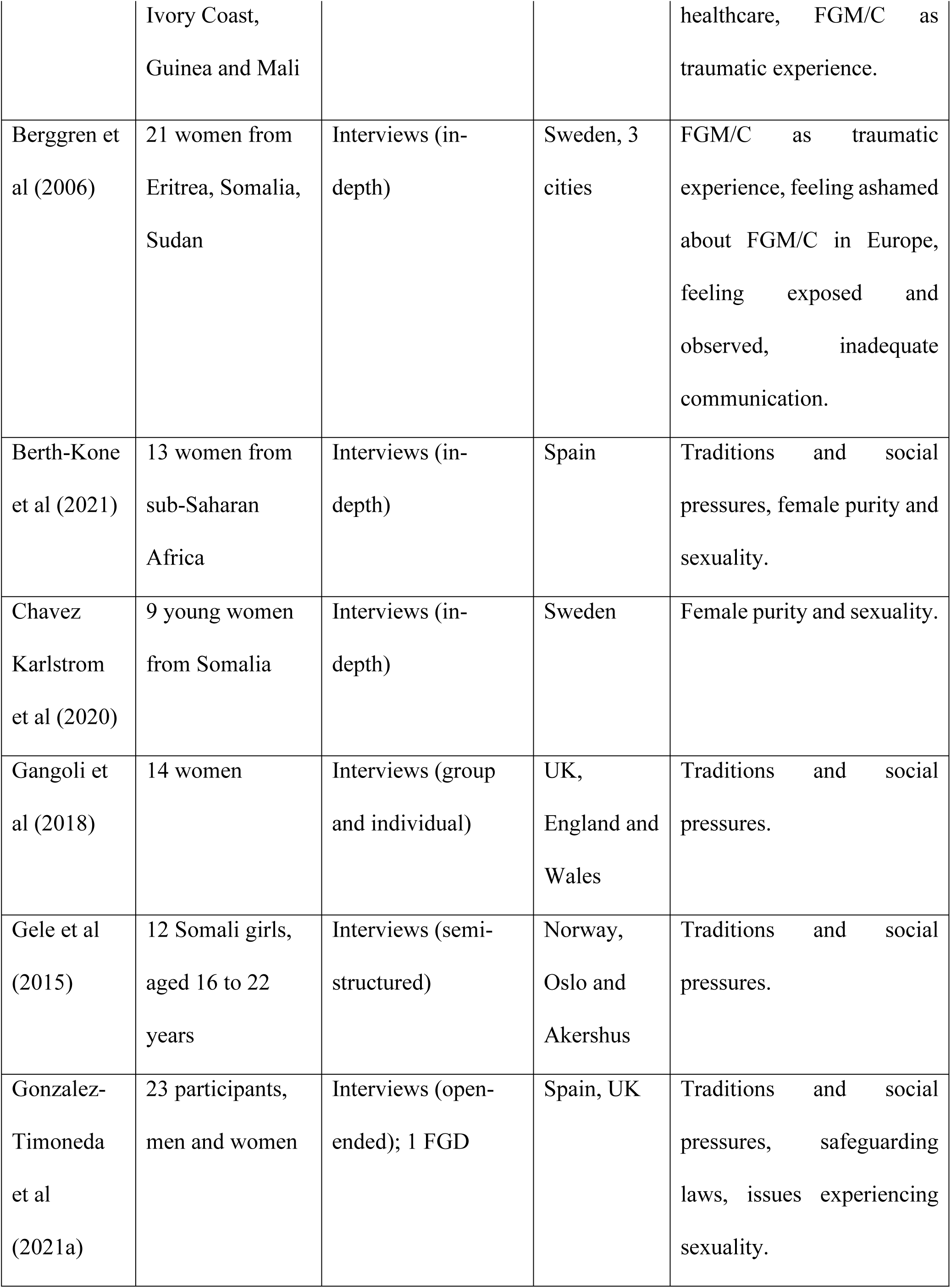
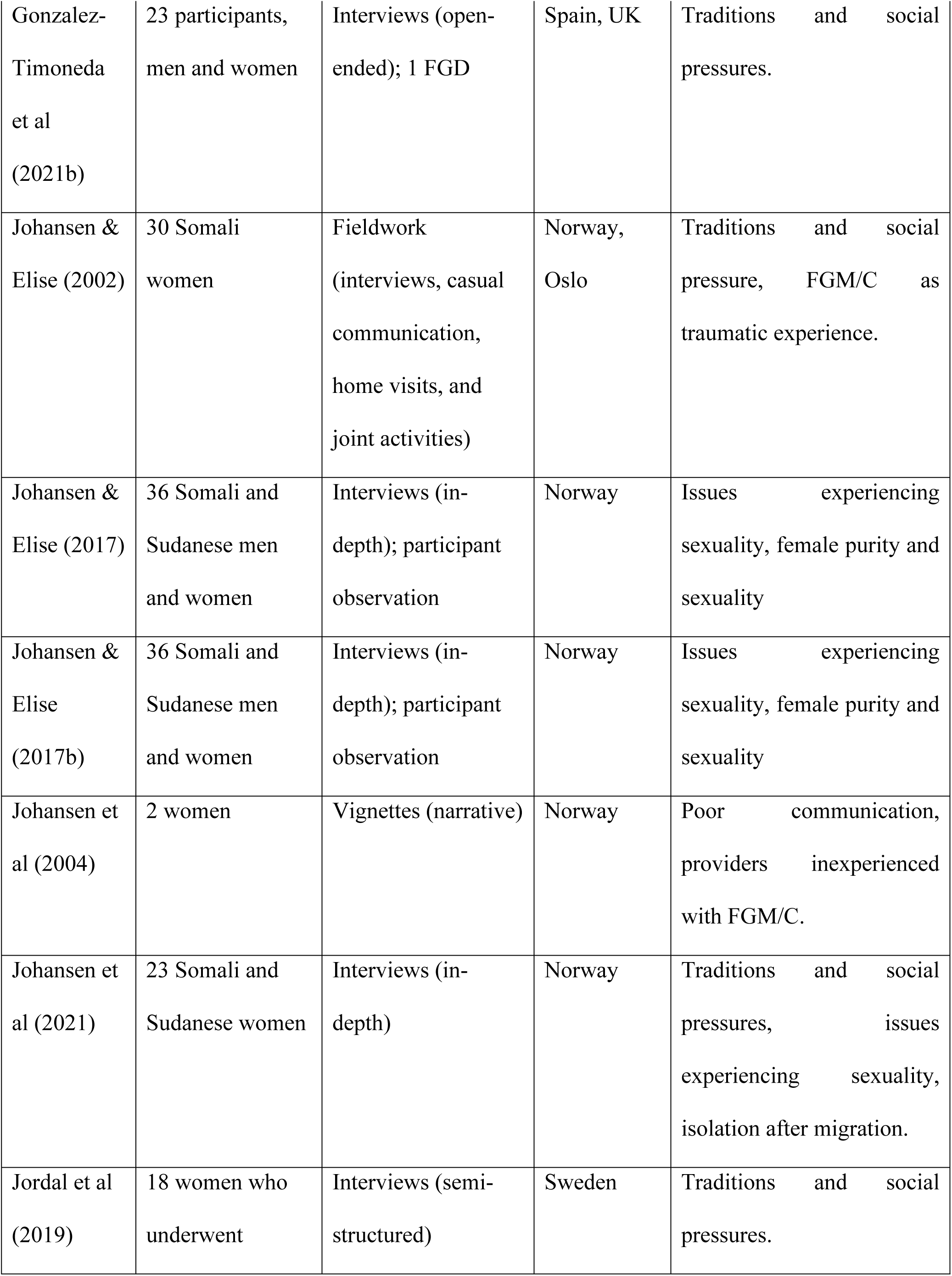
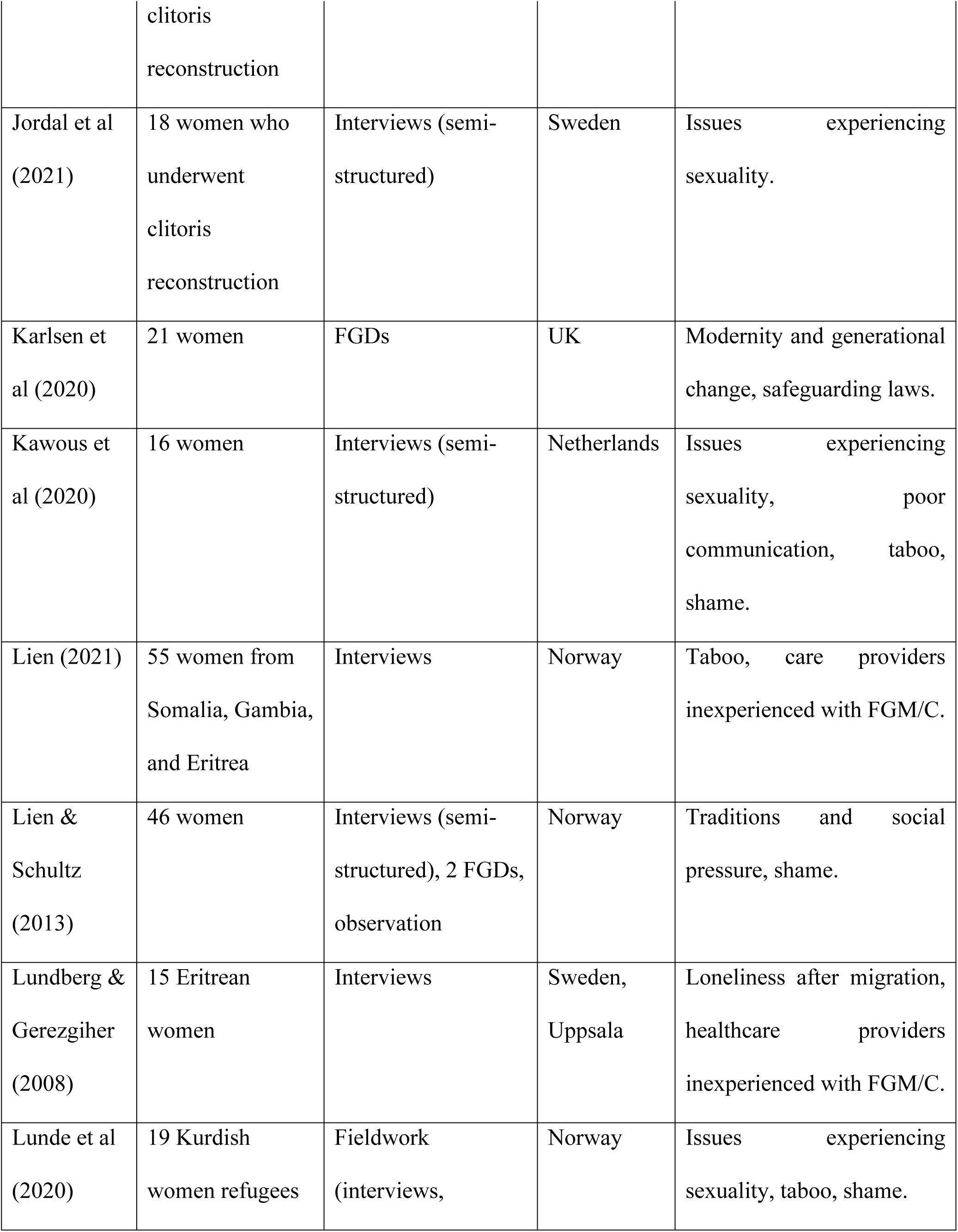
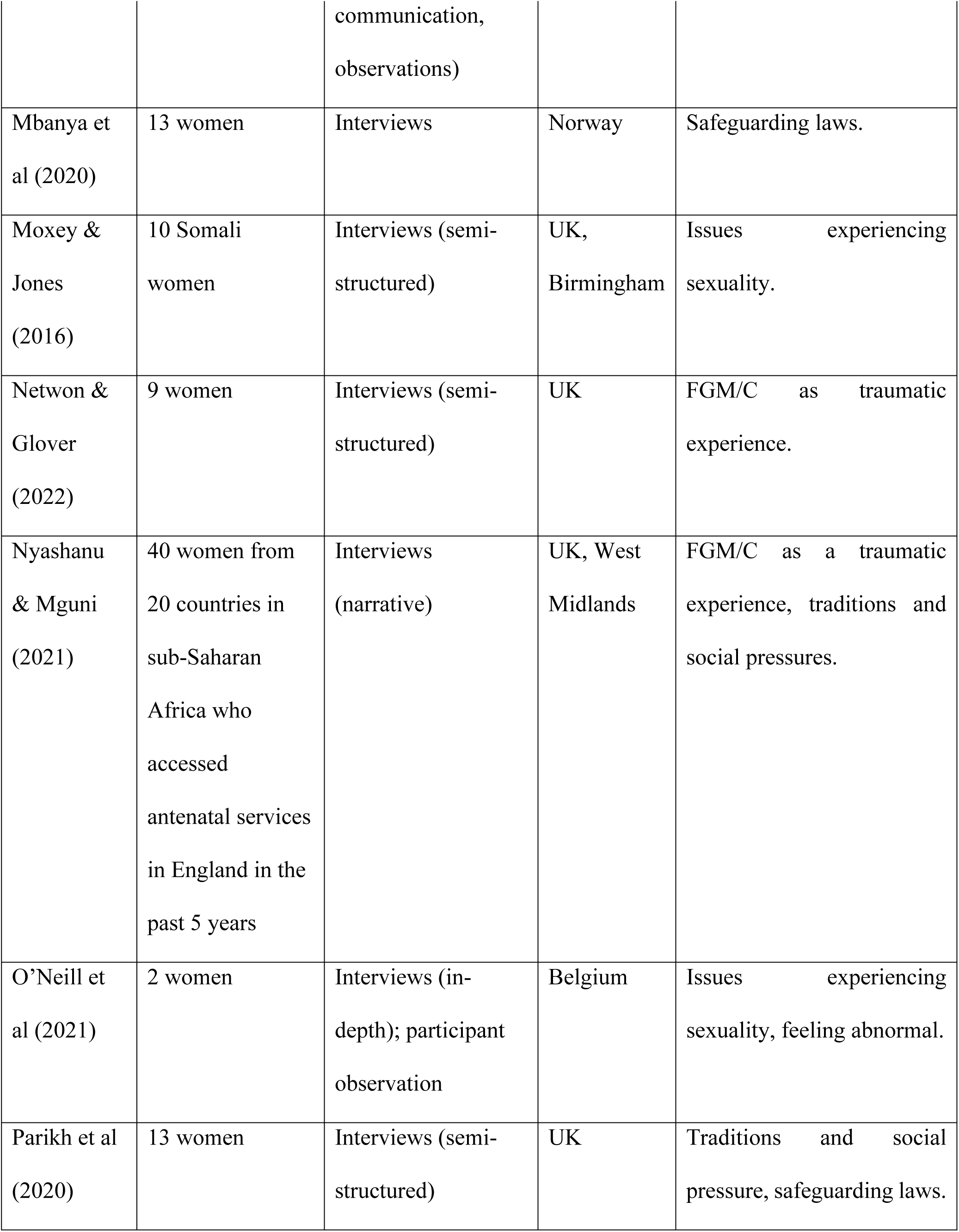
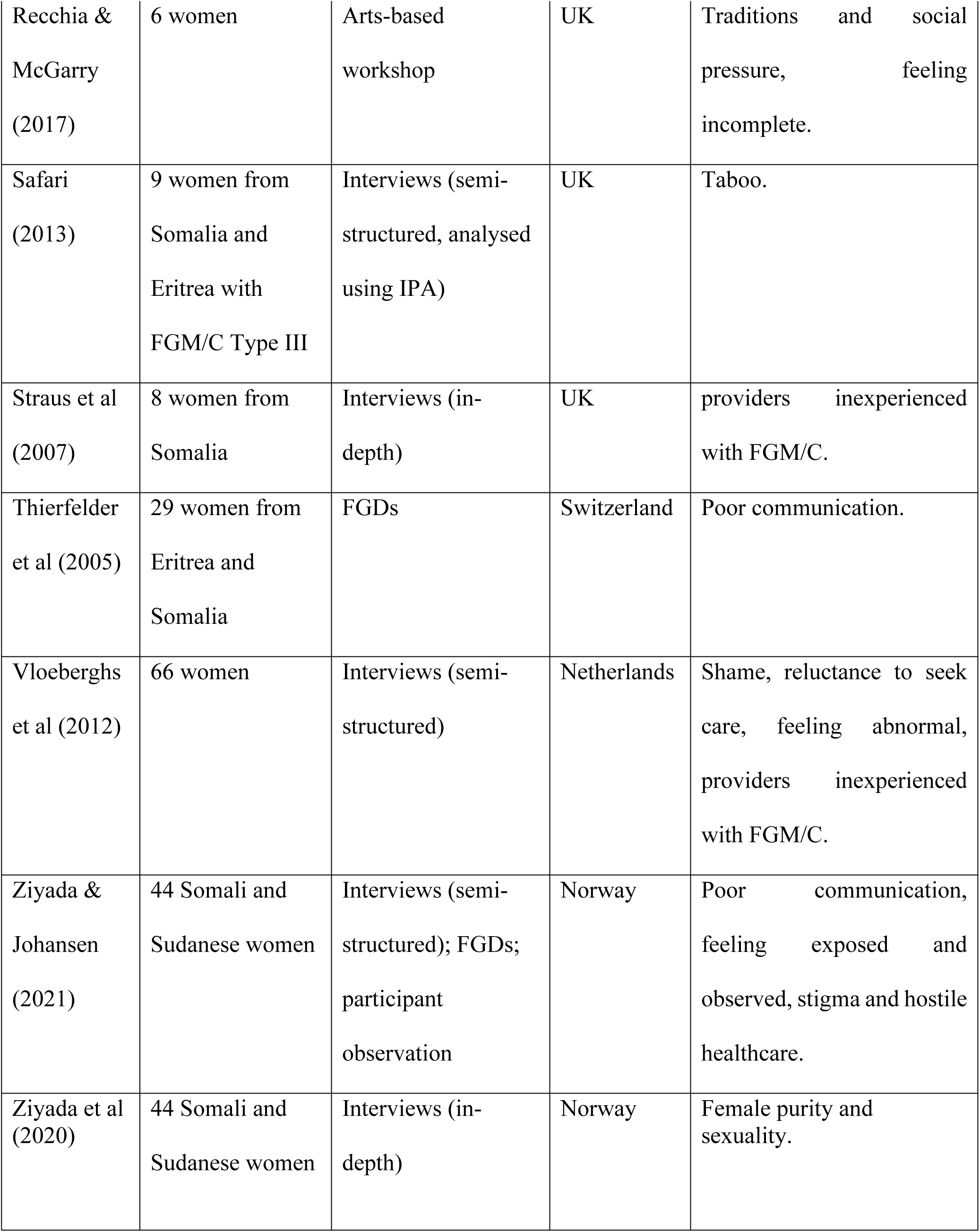
Source characteristics, alphabetically by first author.

### Analytical themes

Following a meta-ethnography approach, we generated three themes that give insight into the experiences of women affected by FGM/C within European healthcare settings, as well as their unique concerns. Themes are: (i) medical ‘othering;’ (ii) challenged communication; and (iii) implications of migration.

#### Medical ‘othering’

This theme refers to women’s descriptions of their emotions when interacting with host country health systems and is derived from accounts linking caregiver actions and attitudes during physical examinations to how women perceived themselves and the quality of care they received. Many women reported feelings of inadequacy, shame, vulnerability, and fear directly linked to feeling different to what healthcare providers might consider ‘normal.’ Interacting with health systems caused women feelings—sometimes internalised—of themselves as foreign, ‘abnormal’, or even ‘incomplete’ for having experienced FGM/C, contrasting with FGM/C being normal or even ‘ideal’ in their countries of origin (Agboli, 2020; Berggren et al., 2006; O’Neill et al., 2022; Recchia & McGarry, 2017; Vloeberghs et al., 2012). In such cases, health systems were experienced as ‘othering’ institutions that actively produced and reminded them of their cultural differences, with implications for the care they received. Providers’ reactions to FGM/C were a key contributor to women’s perceptions of their vulnerability:

> “You can see the facial expressions of the nurse, the doctor, the midwife. You can see their faces, the range of emotions and how they are looking at my body. That hurts…Those people’s eyes make you feel sick.…” (interviewee, Netherlands) (Vloeberghs et al., 2012)

This included experiences that left women feeling like spectacles of medical interest, exposed to observation by multiple staff. For example, being asked if students could enter the room to observe their examination or being collectively stared at by doctors, midwives, and nurses without consent (Berggren et al., 2006; Ziyada & Johansen, 2021) not only evoked feelings of vulnerability, but also of being looked down upon.

> “I would never forget how small I felt at the time. [The gynaecologist] had students in the room. She did not ask me if it was okay. I never agreed to have them staring at my private parts! It was a very uncomfortable situation. I felt violated [and] disrespected.” (interviewee, Norway) (Ziyada & Johansen, 2021)

Additionally, women reported that, despite being subjected to medical scrutiny, they might not receive sufficient attention to their health needs. Instead, many felt judged and even subjected to legal suspicions, especially in countries with safeguarding laws against FGM/C (González-Timoneda et al., 2021; Karlsen et al., 2020; Mbanya et al., 2020; Parikh et al., 2020). This led some to feel re-victimised upon seeking medical care.

> “She [midwife] asked me many questions. She was making as if I have committed a crime.” (interviewee, Norway) (Mbanya et al., 2020)

> “When I finally visited my doctor […] I realized that she was interested to know if my children were circumcised and if I intended to travel with them to Africa. She was not interested in my health needs. When I realized that she was not paying attention to what brought me to the hospital, I immediately left…” (interviewee, Norway) (Mbanya et al., 2020)

#### Challenged communication

This theme explores the difficulties or complete absence of communication between caregivers and affected women. Across most studies, women reported difficulties navigating language and cultural barriers that affected the quality of care they received (Azadi et al., 2022; Berggren et al., 2006; Johansen et al., 2004; Kawous et al., 2020; Ziyada & Johansen, 2021). This challenged communication was due to multiple factors including language, socio-cultural, and gender differences, inappropriate settings, and time constraints. Effective consultation is also dependent on prior knowledge and sensitivity to women’s needs, which is not alleviated solely by having an interpreter (Berggren et al., 2006). The lack of thorough consultation left many women unsatisfied, arriving with the expectation that providers would know how to handle their case and finding the contrary.

> “There are many Somali women here in Sweden, so they must have the experience. This is what I thought. I remember the delivery as a long fight from my side. And then I mean not only fighting with the delivering of my baby but fighting in order to get the staff to understand how I would like it.” (interviewee, Sweden) (Berggren et al., 2006)

Across studies women expressed disappointment in the inadequacy of outreach and support received. Even women who become proficient in their host country’s language/s encountered continuing communication challenges due to usage of complex language and medical terminology by care providers and health educators, and different health promotion culture generally. For example, while host countries focused health promotion efforts on leaflets and written material, many migrant women considered these alien and preferred oral support. Accounts by providers who had experienced FGM/C themselves offered insight:

> “I have all the leaflets here, I translated them, but they are no use at all… it’s oral in our culture…that’s why in my clinic I have to talk to them and talk, talk, talk.” (interviewee, United Kingdom) (Straus et al., 2009)

> “When someone gives me a letter, book, reading no, I don’t want to read it, I want to see and I want to hear it.”(interviewee, United Kingdom) (Straus et al., 2009)

Stigma around FGM/C further challenged communication between affected women and healthcare providers, especially when the latter had limited knowledge of FGM/C (Agboli, 2020; Lien, 2021; Lunde et al., 2021; Safari, 2013). Many women described FGM/C as a topic not openly discussed in their originating cultures, partly because of stigma associated with women’s sexuality. Upon migration, this stigma interacted with stigma derived from the differing norms in their host countries, often translating their experiences with FGM/C into shame. This had tangible consequences in the ways women interacted with health systems, as they felt that interacting with providers would require them to explain things they would prefer not speaking about. Women often felt that the burden was on them to open up and revisit a traumatic experience to receive adequate care, which was attributed as a reason for the reluctance to seek medical services that was common across the studies included. Many women with FGM/C doubted that providers in their host countries would be equipped to deliver care adjusted to their needs, making reluctance to seek care due to distrust, misinformation, and scepticism recurrent events.

> “[FGM/C] is about sex. You cannot share [your FMG/C experiences] with other people. You feel terribly embarrassed. That is why circumcised women become isolated, mentally ill or mad. Either that or she stops talking; she keeps her mouth firmly shut. […] And because we feel ashamed, we stay home with our problems.” (interviewee, Netherlands) (Vloeberghs et al., 2012)

> “My family doctor is a man, and I don’t feel like showing him my private parts. That means having to explain everything all over again, and that is something I absolutely don’t feel like. I don’t want to be reminded of the pain.” (interviewee, Netherlands) (Vloeberghs et al., 2012)

> “I and other women who have not been opened before delivery suffer most. We need to be opened at delivery, but the midwives don’t know how to cut. They wait until the head of the baby is down and then they cut in a hurry in all directions, often several cuttings. They are not careful. I think they see us as already being destroyed.” (Interviewee, Sweden) (Berggren et al., 2006)

#### Implications of migration

This theme developed in relation to most women’s complex circumstances as migrants in addition to living with FGM/C, and how this shaped their healthcare experiences. While we did not restrict eligibility to studies of migrant women, most were first generation immigrants. As migrants, women experienced challenges navigating their new socio-cultural setting while honouring their origins.

Across multiple studies, mothers of young daughters faced social pressures within migrant communities to maintain FGM/C, while other women felt accountable to traditions including FGM/C even without direct social pressures (Alhassan et al., 2016; Berthe-Kone et al., 2021; Gangoli et al., 2018; Gele et al., 2015; González-Timoneda et al., 2022; Johansen & Ahmed, 2021; Lien & Schultz, 2013).

> “If a girl is not circumcised, the other mothers talk about the girl so that the mother hears it and feels ashamed. No mother can stand such talk. You do not want to feel shame for your daughter…”(interviewee, Sweden) (Berggren et al., 2006)

> “Our ancestors did it and they expect us to do it in their absence. They are always watching over us. If you disobey them, it does not matter where you are they can send the punishment to you.” (interviewee, Spain) (Alhassan et al., 2016)

For many migrant women, accessing healthcare in Europe represented the first realisation that not *all* women undergo FGM/C (Agboli, 2020). When this happened, affected women began realising the implications of their experience, often feeling torn between cultural traditions to which they felt accountable and integration within host countries. Many reported concerns that opposing FGM/C could be perceived by their families as abandoning their culture, entailing potential isolation from communities of origin. Conversely, many saw opposing FMG/C as a requirement of integration into their host country, fearing similar stigma and exclusion if they were seen to support or have experienced FGM/C.

> “When you visit a gynaecologist, you are surprised when the doctor tells you that you are not ‘normal’. […] And you realize, after explanation with photos, the difference between the normal and abnormal private part. So, I say, I have never seen the thing between the legs.” (interviewee, Belgium) (Vloeberghs et al., 2012)

> “The first time I saw the genital organ of a woman, I said ah … so I lost this part of me in the excision … But hey, it’s a bit what like I looked as well. But it must be said that this operation is very traumatic. We only perpetuate the tradition of our ancestors. All you gain is pain and sorrow.” (interviewee, Belgium) (Vloeberghs et al., 2012)

> “I would never dare to bring my daughter to Somalia. My grandmother had me cut even when my mother was against it. If I say anything, they say you have a Norwegian tradition.” (interviewee, Norway)(Johansen & Ahmed, 2021)

> “It has happened […] they ask ‘you then, have you, are you also like that, down there? […] So, I said, actually I lied, I said ‘no, I am totally normal’. Yes, in school when they asked, I lied.” (interviewee, Sweden) (Jordal et al., 2019)

Women reported struggles adapting to changes in the forms of available care entailed by migration. Multiple women reminisced about the networks of care they could rely on in their home countries— particularly communities of women—and described a sense of isolation when navigating healthcare in host countries (Johansen & Ahmed, 2021; Lundberg & Gerezgiher, 2008). This was alleviated in some cases when providers demonstrated cultural competence or prior experience working with women living with FGM/C, helping women feel more secure navigating their health needs without the care networks they previously relied on.

> “In my country, women lie on the bed for 40 days [after giving birth] and their movements are limited. They get full support from all members of their families, relatives and even neighbours. One can say that this is the only time they get full rest and good food.” (interviewee, Sweden)(Lundberg & Gerezgiher, 2008)

> “I was lucky when I met a midwife in Sweden who knew about circumcised women. This was a great help to make me feel secure because it was my first time to be pregnant and to live far from parents and family.” (interviewee, Sweden) (Turkmani et al., 2019)

## DISCUSSION

### Key findings

This review, exploring women’s health service experiences in relation to living in Europe with FGM/C, also highlights migration factors that limit or alter affected women’s perceived ability to access healthcare and support. This review is the first qualitative synthesis of healthcare perceptions and experiences of women living with FGM/C in Europe and is also unique in using women’s own narratives through a meta-ethnographic approach across studies. An integral part of ensuring effective health services for affected women is understanding their perceived health needs and experiences while in Europe. Our three themes illustrate women’s feelings about services and describe, in women’s own words, what they still need for healthcare to feel supportive.

Important to our findings, and noted by many scholars, is that the ‘scientific’ knowledge shaping clinical spaces and interactions is neither universal nor objective (Adams, 2001; Good, 1993; Plemons, 2017). Rather, it depends on healthcare provider interpretations of particular models of the body, health, and the meaning of ‘care’, all of which can vary across cultures and individuals (Kleinman, 1997; Scheper-Hughes & Lock, 1987). Clinicians are typically conferred with ‘epistemic authority’, meaning that the forms of knowledge they enact are placed above others and rarely questioned (Popowicz, 2021). Hence, when their way of looking—informed by culturally and individually specific conceptualisations of the female body—produces migrant women living with FGM/C as ‘deviant’, ‘abnormal’, or “already broken”, these views can be easily internalised by women themselves, who may perceive the medical knowledge operating on their bodies to be objective, scientific, and universal. The result is a medical form of ‘othering’, the process by which “boundaries between Us and Them,” always embedded in power relations, are created (Akbulut et al., 2020). For migrant women who are navigating an already-contested integration in Europe beyond the healthcare setting (De Genova, 2017), the medical gaze they experience can become an internalised and embodied marker of non-belonging, beginning with but not limited to feelings of shame and embarrassment. Othering enacted by healthcare providers can intersect between structural violence in care and bordering practices, in which their privileged role in relation to knowledge-bearing can have long lasting consequences in women’s lives (e.g. legal risks for women if providers decide they might cut their daughters).

A need for healthcare capacity to deal with cultural difference and ‘othering’ still appears to exist within the heal systems included. We found that migrant women living with FGM/C struggled across European healthcare settings to access adequate and supportive healthcare. A reason for this was lack of cultural competence and adequate FGM/C knowledge among providers, making their health system interactions potentially re-traumatising. Provider reactions to FGM/C - typically without concealing their astonishment - overlapping with insufficient communication during consultations were primary reasons for this. Consequences for women ranged from reluctance to seek care to inadequate consultations (Koukkula et al., 2016).

### Study implications

Our recommendations reflect the longstanding need for improved cultural competence among clinical and auxiliary health staff and outreach interventions (e.g. community-based health promotion) that foster support between health services, affected women, and broader communities to help women counter the isolation and potential vulnerability they experience navigating host country health systems.

We recommend improved cultural competency training processes for healthcare providers, particularly in countries with significant migrant populations, and feedback mechanisms—anonymous if needed— for migrant women to convey their needs and perspectives on how to enhance their healthcare services. The potential relevance of this recommendation is strengthened by women’s demonstrated willingness to talk about adverse experiences with each other in focus groups or in one-on-one interviews. This indicates their capacity to engage effectively with health services when given a safe avenue to do so. At the core of this recommendation is recognition that the burden should not rest solely on affected women to equip themselves to navigate an often structurally violent health system, but rather to better prepare the system to address their particular needs.

Evidence shows migrant women need adapted health literacy for host health systems, support navigating these systems, and health promotion interventions that empower and provide safe spaces for open dialogue. Migrant support groups can help affected women take ownership of their healthcare experiences by preparing and empowering them to discuss FGM/C with providers prior to any physical examination. This is supported by findings on women’s willingness to discuss their experiences with FGM/C among women with similar experiences.

Though FGM/C is described as a complex socio-cultural and healthcare issue (Abdulcadir et al., 2011), interventions addressing FGM/C within potentially-affected diaspora communities in Europe have focused on FGM/C elimination efforts, often ignoring the circumstances of migrant women living with FGM/C. Appropriate services provision for affected women is crucial to addressing FGM/C in Europe (Baillot et al., 2018), yet implications of ways migrant women living with FGM/C describe navigating host health systems (e.g. embarrassment/shame, reluctance to access services altogether) indicate more needs to be done. Reasons for reluctance appear rooted in lack of support, lack of a female community, challenging communication, attitudes of healthcare providers, and (reasonable) distrust of providers’ knowledge and skills in de- and re-infibulation. Advocacy and support networks for women living with FGM/C within diaspora communities can help affected women support one another to navigate challenging host health systems.

### Limitations

Several limitations should be considered. First, coverage across greater Europe was limited to a few countries, with Central, Eastern, and Southeastern Europe entirely absent. More primary research is needed. Second, eligible studies relied on qualitative data collection methods, primarily in interviews and focus group discussions, which may have limited participation to women more able and willing to actively engage. Third, Noblit and Hare’s meta-ethnography only offers a vague approach to qualitative synthesis, as others have noted (Sattar et al., 2021). Additionally, meta-ethnography is normally conducted on systematic review data. However, we found that it worked equally well with our broader scoping review data. Finally, few co-authors’ personal and cultural backgrounds are directly shaped by FGM/C, limiting our lived experience of FGM/C.

## Conclusion

Women living with FGM/C face significant, often unintended, barriers when accessing healthcare services in European countries, this can be particularly challenging for migrant women and impact their ability to thrive in a new social environment. Interventions have focused on eliminating FGM/C, often overlooking the challenges affected women experience. Reluctance to access services can arise from challenged communication, lack of female networks, unsupportive healthcare, and discomfort with providers. Comparing European healthcare to that in their home countries, women expressed a need for more community support. Above all, our findings indicate the need to acknowledge and redirect the power of medical providers, who can inadvertently act as agents of cultural bordering when interacting with migrant women.

## Data Availability

All data is available online.

## DECLARATIONS

### Author contributions

SH and NH conceived the study. SH and MM and developed search terms and syntax with support from NH. All authors contributed to screening and extracting data. SH and MDLT drafted the manuscript with contributions from CN and AH. NH critically revised the manuscript. All authors approved the version for submission.

## Acknowledgements

We are grateful to Toh Kim Kee and Stephanie Ng Yen Ping at the National University of Singapore Library, and Kate Perris at the LSHTM Library for helping develop search syntax.

## Data availability

Original literature sources are all available online.

